# The development and usability testing of two digital knowledge translation tools for parents of children with urinary tract infections

**DOI:** 10.1101/2021.06.21.21259281

**Authors:** Anne Le, Lisa Hartling, Shannon D. Scott

## Abstract

Urinary tract infections (UTI) are a common source of acute illness for infants and children. Approximately 7-8% of girls and 2% of boys will experience a UTI before they are 8 years old. UTIs may be difficult to identify and treat as symptoms in children are different from expected adult symptoms. A previously conducted systematic review identified four common information needs expressed by parents. More specifically, the research identified that parents had difficulty recognizing signs and symptoms of UTIs, felt disappointed by health care provider’s responses, needed timely and relevant information, and feared the unknown due to lack of UTI knowledge. This demonstrates that more effective knowledge translation tools are needed to satisfy parent information needs.

The purpose of this research was to work with parents to develop and test the usability of an interactive infographic and video about UTIs in children. Prototypes were evaluated by parents through usability testing in two Alberta emergency department waiting rooms. Results were positive and overall, the tools were highly rated across all usability items, suggesting that arts-based digital tools are useful mediums for sharing health information with parents.

## Introduction

Urinary tract infections (UTI) are a common source of acute illnesses for infants and children [1]. About 7-8% of girls and 2% of boys will experience a UTI before they are 8 years old [2]. Most children will recover without complications with proper antimicrobial therapy [3]. Some children, however, may develop lifelong consequences such as hypertension, decreased renal function, and proteinuria due to the renal scarring caused by UTIs [1-2]. Risk for complications is especially high if UTIs are not diagnosed and treated appropriately [1,4]. UTIs in children may be difficult to treat and identify as symptoms typically different from adult symptoms [3,5]. In addition, there are wide variations in diagnosis methods and treatments for pediatric UTIs which may compound to increase confusion for parents, delaying care [1,6]. It is critical that parents be provided with accurate information on diagnosis and treatment options that will facilitate educated decision-making regarding their child’s care.

A previously conducted systematic review identified four common information needs expressed by parents [8]. More specifically, the research identified that parents had difficulty recognizing signs and symptoms of UTIs, felt disappointed by health care provider’s responses, needed timely and relevant information, and feared the unknown due to lack of UTI knowledge [8-9]. This demonstrates that more effective knowledge translation tools are needed to satisfy parent information needs [7]. Evidence has suggested that developing KT tools to target health consumers, such as caregivers, may aid decision making and understanding of treatments [10,11].

Research exploring the benefits of art and narrative based forms of KT tools have illustrated the power these forms may have in communicating, engaging, and influencing individuals [11-16]. Currently, there have been limited numbers of KT tools developed for UTI education. Tools that currently exist target different populations, such as pregnant women and seniors, are for use by healthcare providers (HCP), or are meant to be used along with HCP education sessions. To date, there have been little to no tools developed for UTI education targeted specifically for pediatric and parent populations using an art and narrative form on a digital platform. The purpose of this research was to work with parents to develop and assess the usability of an interactive infographic and video about UTIs in children.

## Methods

We employed a multi-method study involving patient engagement to develop, refine, and evaluate a whiteboard animation video and interactive infographic for pediatric UTIs. Research ethics approval was obtained from the University of Alberta Health Research Ethics Board (Edmonton, AB) [Pro00062904]. Operational approvals were obtained from individual emergency departments and urgent care centres to conduct usability testing.

### Compilation of Parents’ Narratives

Parental narratives were informed through semi-structured qualitative interviews (**Appendix A**) and a systematic review [17-18]. Parents of children who presented to the Stollery Children’s Hospital (Edmonton, Canada) with urinary tract infections were invited to participate. Interviews (n = 18) were conducted by a research coordinator trained in qualitative methodology. Parents were asked to share their experiences having a child with UTI. Concurrently, a systematic review was conducted to synthesize current evidence about experiences and information needs of parents managing UTI. Results from the systematic review and qualitative interviews are published elsewhere [17-18].

### Intervention Development

Using the key findings from the systematic review and qualitative studies, researchers developed an infographic skeleton and video script, which included all information that was to be integrated into the tools. This included key quotes and salient themes from the compilation of parents’ narratives and recommendations from the TRanslating Emergency Knowledge for Kids (TREKK) Bottom Line Recommendations (BLR) for UTI in children [19]. Creative writers, illustrators, graphic designers, and videographers were selected using a competitive process. The video storyline depicted parents struggling to manage their children’s condition, along with the necessary information on how to care for their child and when to seek professional help (**Appendix B**). Similarly, information on what causes UTIs, symptoms, treatment options, and when to seek emergency care were included in the infographic (**Appendix C**).

### Revisions

Iterative processes were used to develop the tools. Different stakeholder groups, such as parents, HCPs, researchers, and the study team provided significant feedback on the tools. HCPs were asked to comment on the clinical accuracy of information and evidence, usefulness, and perceived value, while parents from our Pediatric Parent Advisory Group (P-PAG) were asked to provide feedback on the length, stylistic elements, and highlight areas/information needs not addressed in the tools. The P-PAG, a group launched by the principal investigators of ECHO and ARCHE, meets once a month and is asked to provide input on KT tools and other research activities. Likewise, research team meetings are held weekly to discuss the development of our tools. Feedback from these meetings is aggregated and provided to developers for revisions. A visual depiction of our KT Tool Development processes is displayed in **Figure 1**.

**Figure 1.**
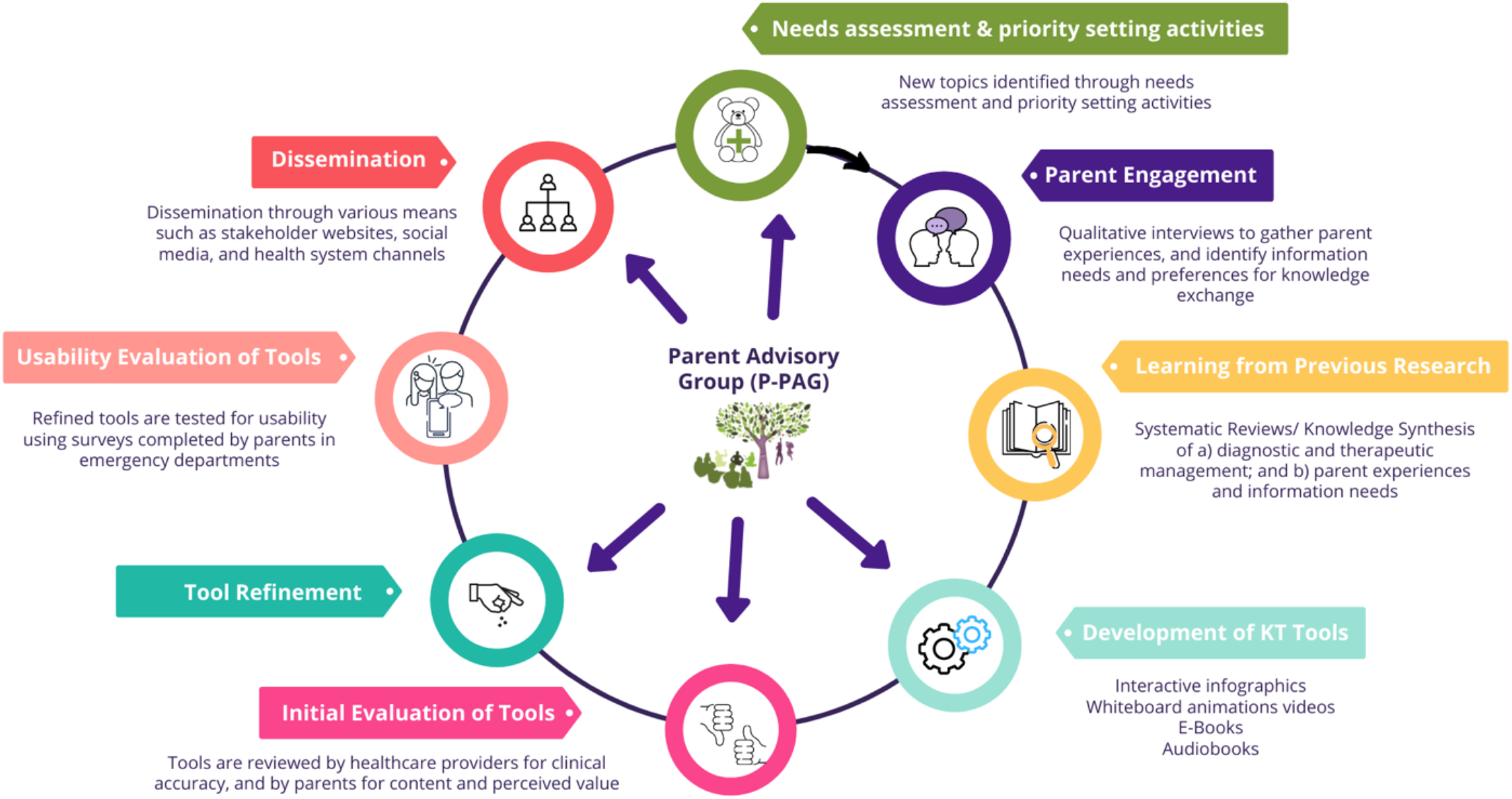
ARCHE-ECHO Tool Development Cycle.

### Video

Five versions of the video were developed prior to usability testing. The English-language, close captioned video that was used for usability testing depicted a mother’s experiences when her baby develops a UTI. In 5 minutes and 38 seconds, the video discusses the symptoms that may occur with a UTI and when a parent should take their child to the emergency department (ED), family doctor, or walk in clinic for treatment. It illustrates the types of diagnostic tests performed, the variance in symptoms based on age, followed by common treatments. It concludes with tips to prevent UTIs and what to expect following treatment.

### Infographic

The UTI interactive infographic was developed using the same style, design elements, and animations used in our suite of infographics. The style is unique to our research program and developed over the course of two years.

The infographic functions similarly to a webpage and allows users to scroll through the information, exploring at their own pace. The information provided in the UTI infographic mirrors information provided in the video. In total, the infographic included 7 sections: (1) What is a UTI?, (2) UTI Symptoms, (3) When to Seek Care, (4) Treatment, (5) Useful Links, and (6) Contact Us. Under UTI Symptoms, the experiences of parents with children affected by UTIs are added to demonstrate the variety of symptoms that may occur.

### Surveys

Parents were recruited to participate in an electronic, usability survey (**Appendix D**) in two urban ED waiting rooms in the Edmonton area (Stollery Children’s Hospital and Northeast Community Health Centre). Surveys were informed by a systematic review of over 180 usability evaluations and comprised of 9, 5-point Likert items assessing: 1) usefulness, 2) aesthetics, 3) length, 4) relevance, and 5) future use [20]. Parents were also asked to provide their positive and negative opinions of the tool via two free text boxes. Members of the study team approached parents in the ED to determine interest and study eligibility. Study team members were available in the ED to provide technical assistance and answer questions as parents were completing the surveys.

### Data Analysis

Data were cleaned and analyzed using SPSS v.24. Descriptive statistics and measures of central tendency were generated for demographic questions. Likert answers were given a corresponding numerical score from 5 to 1, with 5 being “Strongly Agree” and 1 being “Strongly Disagree” [21-22]. T-tests were conducted to determine whether there were any significant differences between usability means for the two tools. Open-ended survey data were analyzed thematically. A summary of the results was then shared with the creative team to inform the development of the final versions of the tools.

## Results

Usability of the UTI infographic and video was evaluated using 9 questions and 2 free text boxes. Each question was rated from *strongly disagree* (1) to *strongly agree* (5). 61 parents awaiting pediatric ED care were approached to complete a usability survey. 30 complete responses were submitted for both infographic and video usability testing.

Parental reaction to the infographic and video were generally positive as all mean results were between 4 (*agree*) and 5 (*strongly agree*). Usability testing illustrated that parents generally believed the video was more useful than the infographic in conveying UTI information. The mean response for infographic usefulness was 4.10 whereas a mean of 4.77 for the video (p = 0.005). Although the differences in results for the remaining questions were not significant, slightly more parents believed the video was more relevant, easier to use, and aesthetically pleasing than the infographic whereas a few more parents believed the infographic had a better length and would be more useful in the future and when making decisions on their child’s health. When asked if the tool was relevant to parents, a mean response of 4.27 was generated for the infographic and 4.45 for the video. Moreover, a mean response of 4.48 was recorded for infographic simplicity whereas the video recorded a mean of 4.63. The mean answer, when asked about the ability to use the infographic without additional help, was 4.40 with a mean of 4.62 for the video. A mean result of 4.37 was generated for the infographic length and a mean of 4.17 for the video length. The results for aesthetic yielded a mean of 4.43 for the infographic and 4.48 for the video. When asked if the tool would be used in the future, a mean of 4.33 was recorded for the infographic and a 4.07 mean for the video. Likewise, when parents were asked if the tool would help make decisions about their child’s health, a mean answer of 4.17 resulted for the infographic and a mean answer of 4.10 for the video. A mean answer of 4.30 resulted when parents were asked if they would recommend the infographic to a friend and a mean answer of 4.47 was generated when asked likewise about the video. **Table 2** displays the mean responses to each question for each tool.

**Table 1.** Demographic characteristics of parents who assessed the usability of the UTI e-tools (N=60)

| Characteristic | n (%) |
| --- | --- |
| Sex |  |
| Female | 45 (75.0) |
| Male | 14 (23.3) |
| Missing | 1 (1.7) |
| Age |  |
| Less than 20 | 1 (1.7) |
| 20-29 years | 10 (16.7) |
| 30-39 years | 22 (36.7) |
| 40-49 years | 15 (25.0) |
| 50-59 years | 5 (8.3) |
| 60 years and older | 7 (11.7) |
| Marital Status |  |
| Married/Partnered | 42 (70.0) |
| Single/Separated/Divorced/Widowed | 18 (30.0) |
| Education |  |
| Completed primary school | 1 (1.7) |
| Some high school | 5 (8.3) |
| High school diploma | 8 (13.3) |
| Some post-secondary | 6 (10.0) |
| Post-secondary certificate/diploma | 15 (25.0) |
| Post-secondary degree | 15 (25.0) |
| Graduate degree | 8 (13.3) |
| Other | 2 (3.3) |
| Household Income |  |
| Less than \$25,000 | 4 (6.7) |
| \$25,000-\$49,000 | 10 (16.7) |
| \$50,000-\$74,000 | 11 (18.3) |
| \$75,000-\$99,000 | 12 (20.0) |
| \$100,000-\$149,000 | 7 (11.7) |
| \$150,000 and over | 9 (15.0) |
| Prefer not to answer | 4 (6.7) |
| Missing | 3 (5.0) |

| Number of Children in the Family |  |
| --- | --- |
| 1 | 17 (28.3) |
| 2 | 19 (31.7) |
| 3 | 17 (28.3) |
| 4 | 5 (8.3) |
| 5+ | 2 (3.3) |

**Table 2.** Means (SD) of participant responses to the usability survey.

| Usability Measures | Video | Infographic |
| --- | --- | --- |
| It is useful. | 4.77 (0.43) | 4.10 (1.16)* |
| It provides information that is relevant to me as a parent. | 4.45 (0.74) | 4.27 (0.83) |
| It is simple to use. | 4.63 (0.56) | 4.48 (0.87) |
| I can use it without written instructions or additional help. | 4.62 (0.49) | 4.40 (0.93) |
| Its length is appropriate. | 4.17 (0.70) | 4.37 (0.81) |
| It is aesthetically pleasing (i.e. images, colours, etc.). | 4.48 (0.58) | 4.43 (0.73) |
| It helps me make decisions about my child's health. | 4.07 (0.74) | 4.33 (0.76) |
| I would use it in the future. | 4.10 (0.66) | 4.17 (0.79) |
| I would recommend it to a friend. | 4.47 (0.73) | 4.30 (0.65) |

\*p < 0.05.

While there were limited comments in the open text boxes, parents only had positive things to say about the UTI video. Parents described the UTI video as being “very well done” and encouraged researchers to “continue doing this kind of research more often.”

There were more comments pertaining to the UTI infographic, both negative and positive. A majority of the comments were positive and centred mainly around the aesthetics of the tool and its visual appeal. Parents said that they made the tool “easy to understand” and more “attractive”. They also said that the images helped improve their knowledge of UTIs. In terms of negative comments, parents felt that the tool may be too repetitive.

## Conclusions

The purpose of this project was to develop two arts-based digital tools (a whiteboard animation video and interactive infographic) for parents of children who have urinary tract infections. Using a multi-method approach that encompassed stakeholder engagement, we were able to create tools that were highly rated amongst parents seeking care for their children in two urban emergency care centres. Parents found the tools to be useful, relevant, easy to use, and aesthetically pleasing. Most importantly, parents felt that the tools could facilitate decision-making in the future, with parents mainly agreeing and strongly agreeing that they would use the tool and recommend the tool to their friends. Results from this project indicate that end-user engagement plays a positive role in developing knowledge tools that address the needs of parents.

**The tools can be found here: http://www.echokt.ca/tools/urinary-tract/**

Note: Our KT tools are assessed for alignment with current, best-available evidence every two years. If recommendations have changed, appropriate modifications are made to our tools to ensure that they are up-to-date [23].

## Data Availability

Data referred to in the manuscript are not available for access.

## Author Contributions

This study was conducted under the supervision of Drs. Shannon D. Scott (SDS) and Lisa Hartling (LH), PIs for **translation Evidence in Child Health to enhance Outcomes** (ECHO) Research and the **Alberta Research Centre for Health Evidence** (ARCHE), respectively. Both PIs designed the research study and obtained research funding through Translating Emergency Knowledge for Kids (TREKK) Networks of Centres of Excellence of Canada (NCE), Stollery Children’s Hospital Foundation, and the Women and Children’s Health Research Institute.

SDS designed and supervised all aspects of tool development and evaluation.

LH contributed to tool development and evaluation and supervised the systematic review of parent experiences and information needs.

Alyson Campbell and Samantha Louie-Poon collected and analyzed qualitative interviews with parents.

Tony An developed the infographic.

Tabatha Plesuk and Hyelin Sung collected usability data.

Anne Le analyzed usability data.

All authors contributed to the writing of this technical report and provided substantial feedback.

This work was funded by:

**Networks of Centres of Excellence:**

- Klassen, T., Hartling, L., Jabbour, M., Johnson, D., & Scott, S.D. (2015). Translating emergency knowledge for kids (TREKK). Networks of Centres of Excellence of Canada Knowledge Mobilization Renewal ($1,200,000). January 2016 – December 2019.

**Women’s and Children Health Research Institute**

- Scott, S.D & Hartling L. (2016). Translating Emergency Knowledge for Kids renewal. Women and Children’s Health Research Institute (matched dollars, $150,000). 2016/04-2019/04.

## Other Outputs from this Project

## Appendices

### Appendix A Interview Guide

Parents will be interviewed to understand their experience having a child with a UTI. Semi-structured interviews will be conducted with parents in order to get their “narrative” or experiences. The following questions will be used to guide these interviews. Being true to semi-structured interview techniques, interview questions will start broad and then move to the more specific.

1. Tell me about your experience having your child experience a UTI.
  a. What were the symptoms? How was your child behaving? How did you know they were sick? Has this happened before?
  b. When did you decide to take them to the emergency department (ED)? Why did you decide to go to the ED?
  c. Did your child’s illness affect your day to day activities? Were you getting the usual amount of sleep? How did it affect your family? (partner other kids)
2. Tell me about your child that was ill.
  a. How old is your child? How was your child ill? How were they feeling? Describe this to me. Were they ‘out of sorts’? Were they eating and sleeping as they normally would? Did they miss their usual activities?
  b. Has your child previously had a UTI? If so, how many times? How was this time different? Was it different?
3. Can you tell me about any thoughts or feelings you were experiencing during this time?
  a. Were you worried, scared, nervous? Was it stressful? If so, how was it stressful?
4. Tell me how prepared you felt during the experience.
  a. Did you feel confident in what to do to care for your child? Were you confident that you made the right choice to go to the ED? Did you go to your doctor or call the doctor before going to the ED? Did you call or talk to anyone else to seek advice, such as a friend? Family member? Other health care professional?
5. Did you have all of the information you needed to make decisions about when to seek healthcare? Tell me more about that.
  a. Did you look for information when your child first became ill? Did you look for information about whether or not to go to the ED or see a doctor?
  b. Where did you find information? What did you find? Did you find anything that was helpful? If so what was it and where did it come from?
  c. Where would you typically look for health information for your child? Have you found information in the past? If so, what type of information was it and was it helpful? What do you think would makes information useful?
6. Do you use social media?
  a. If so, did you look for information or ask questions on social media? Would you use social media as a place to get or ask for health information for your child? Which social media platforms do you use (Facebook, Instagram, snapchat, other).
7. What did you do to manage symptoms of UTI? (any techniques you used, for example, giving Tylenol, talking with family/friends, etc.)
  a. How did you feel about your treatment regime? Did you feel confident in what you did? How did your child respond to this?
8. How was your experience in the ED? Tell me about it.
  a. Did you have to wait very long? Approximately how long? How were you feeling during your wait? How was your child feeling? Did you find things for your child to do while waiting? Tell me about your interactions with the healthcare team. Was it an overall positive experience? If so, what made the experience positive? If not, what made the experience negative?
9. Tell me about when or how your child was diagnosed with a UTI – were any tests done? Any medications ordered?
  a. What tests were done? Were these tests explained to you and why they were necessary? Were you uncomfortable with any of the tests that were done (blood work, ultrasounds, other)? If so, what made you uncomfortable about them? How was your child during these tests - were they nervous, anxious, crying, etc? Do you feel you got all the information you needed about what was happening?
10. What strategies were put in place by health care professionals to help your child? (for example, giving/prescribing medication). Did they ask you to do anything? If so, how comfortable were you with that? Did they ask you what you have already tried?
  a. Did they give you any information before you went home from the ED? If yes, what did they give you? Did they give you any advice for what to do at home? When to see your doctor? or when to come back to the ED?
11. How did your child manage the experience? How did you feel about the outcome of this situation? Did it go as expected for you?
  a. Was your child anxious, nervous? Did everything go as you had hoped or planned? Did you have any follow up – other tests, going back to the doctor?
12. If presented with the same situation again (your child being ill with a UTI), would you do anything differently? If so, please tell me.
  a. How would you make a decision about whether or not to go to the ED? Would you look for information before going this time? Where would you look or who would you ask for advice?
13. If the health system were to have information for parents, what do you think would be the best way to get it to parents? Through their website, call line, advertisements, social media, public health clinics, doctors’ offices, etc.?

### Appendix B Video Images

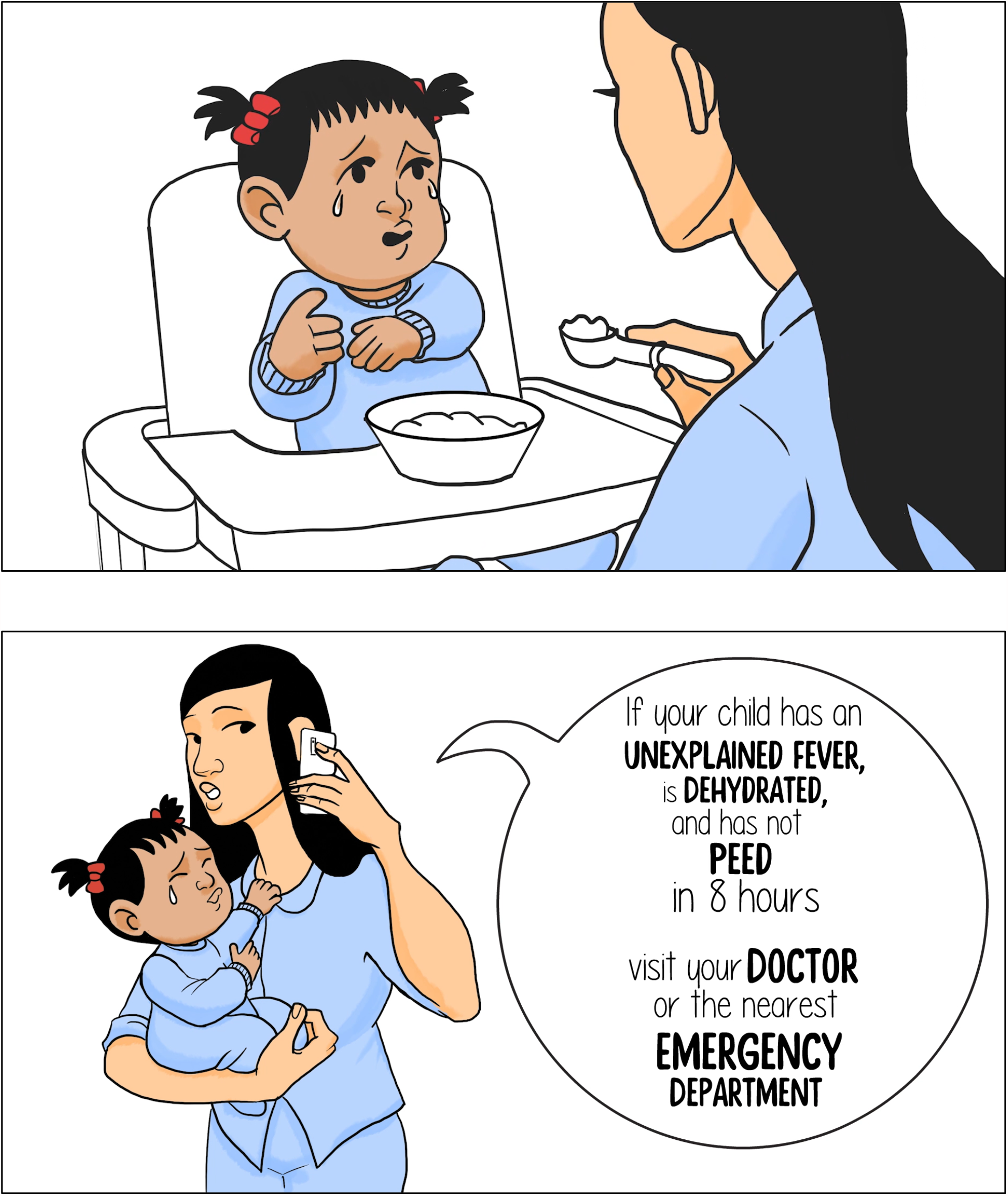

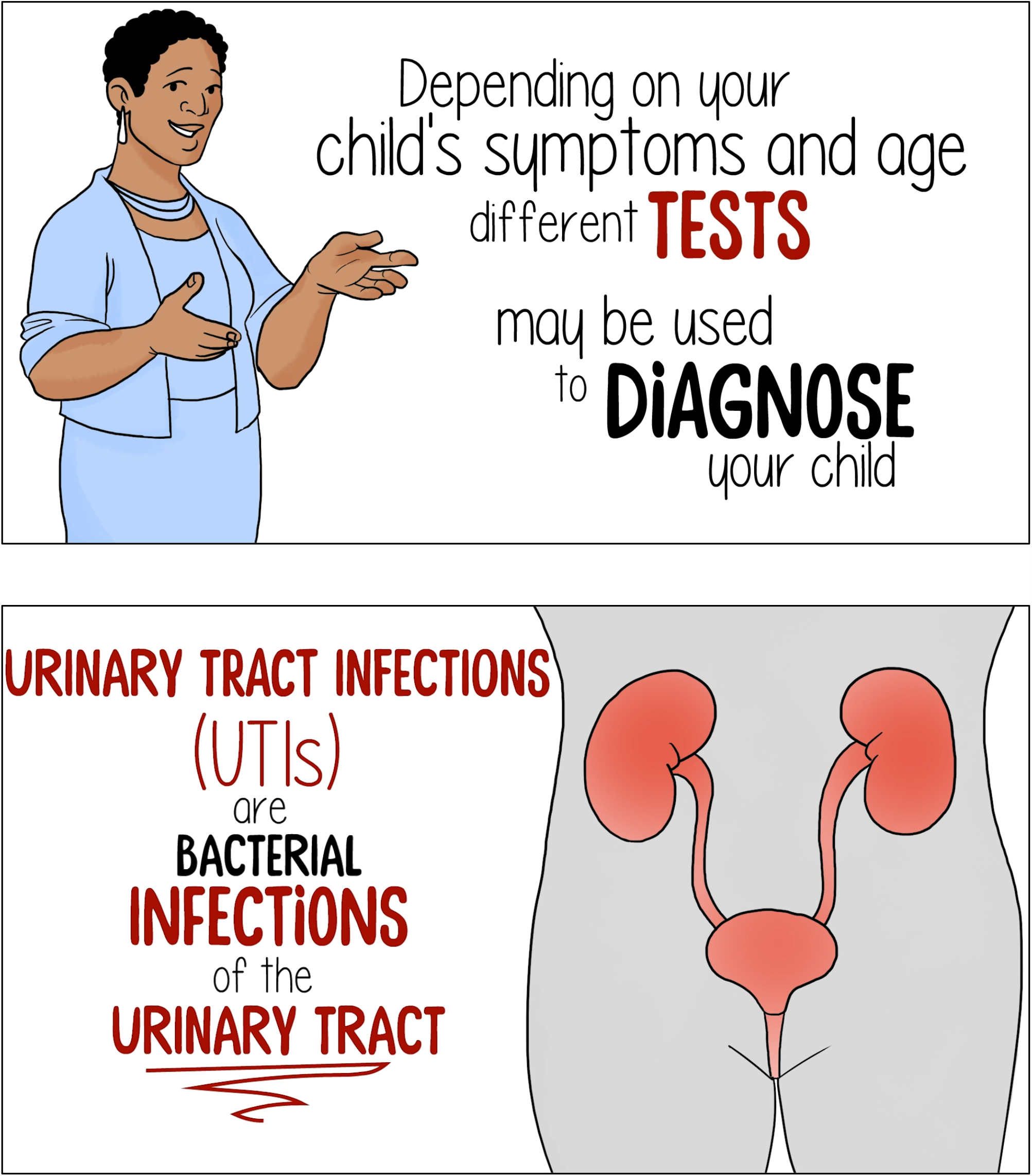

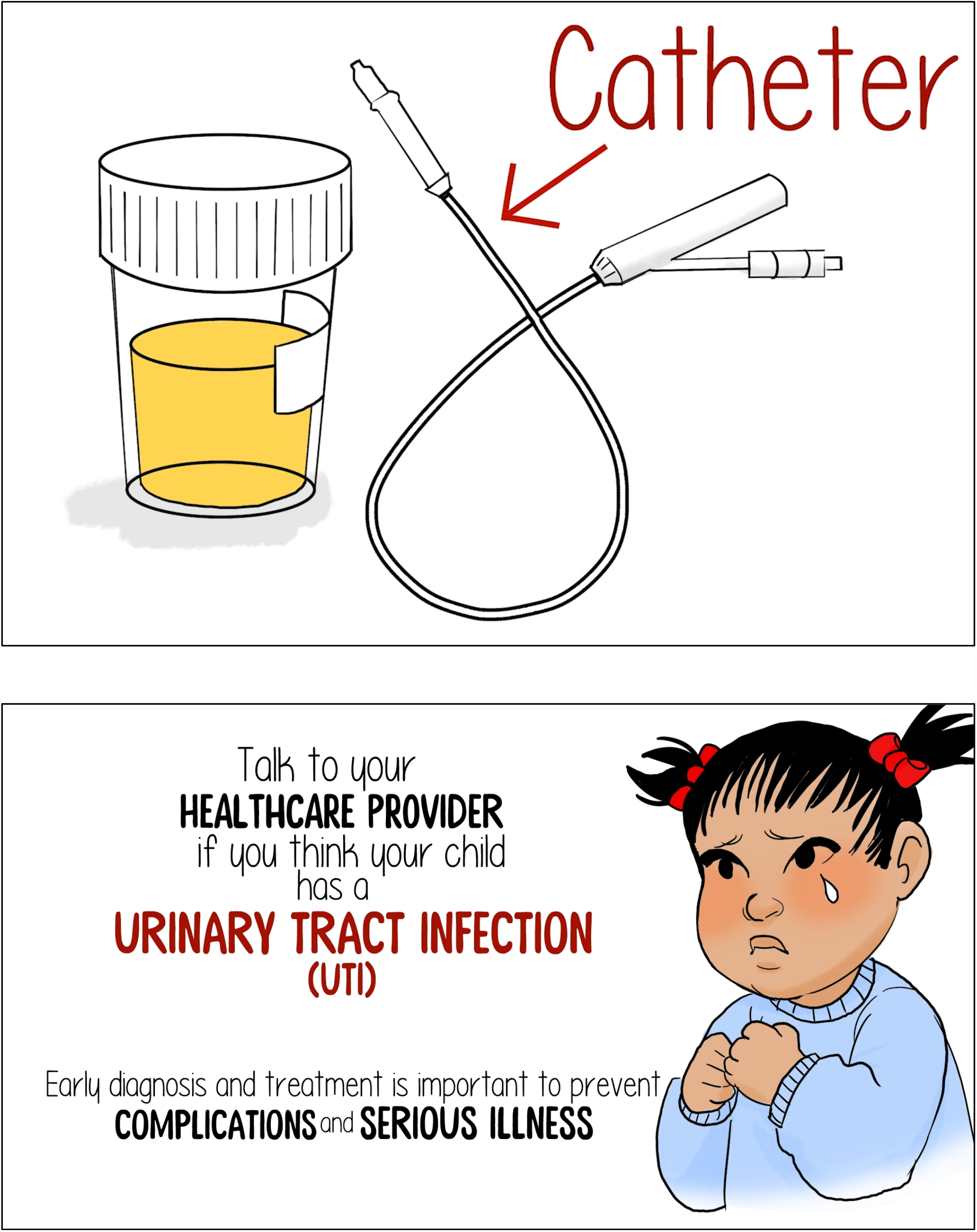

### Appendix C Infographic Images

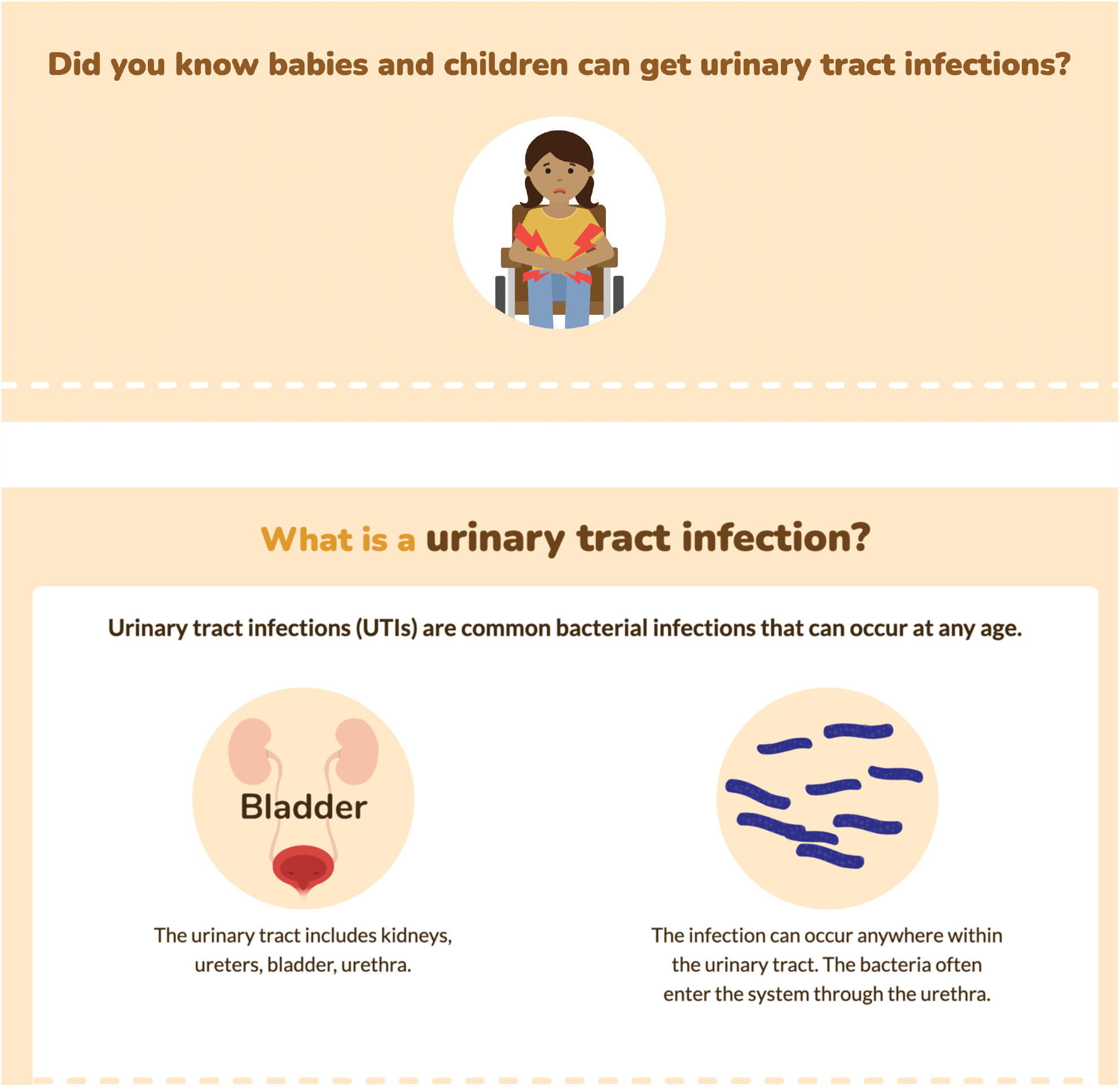

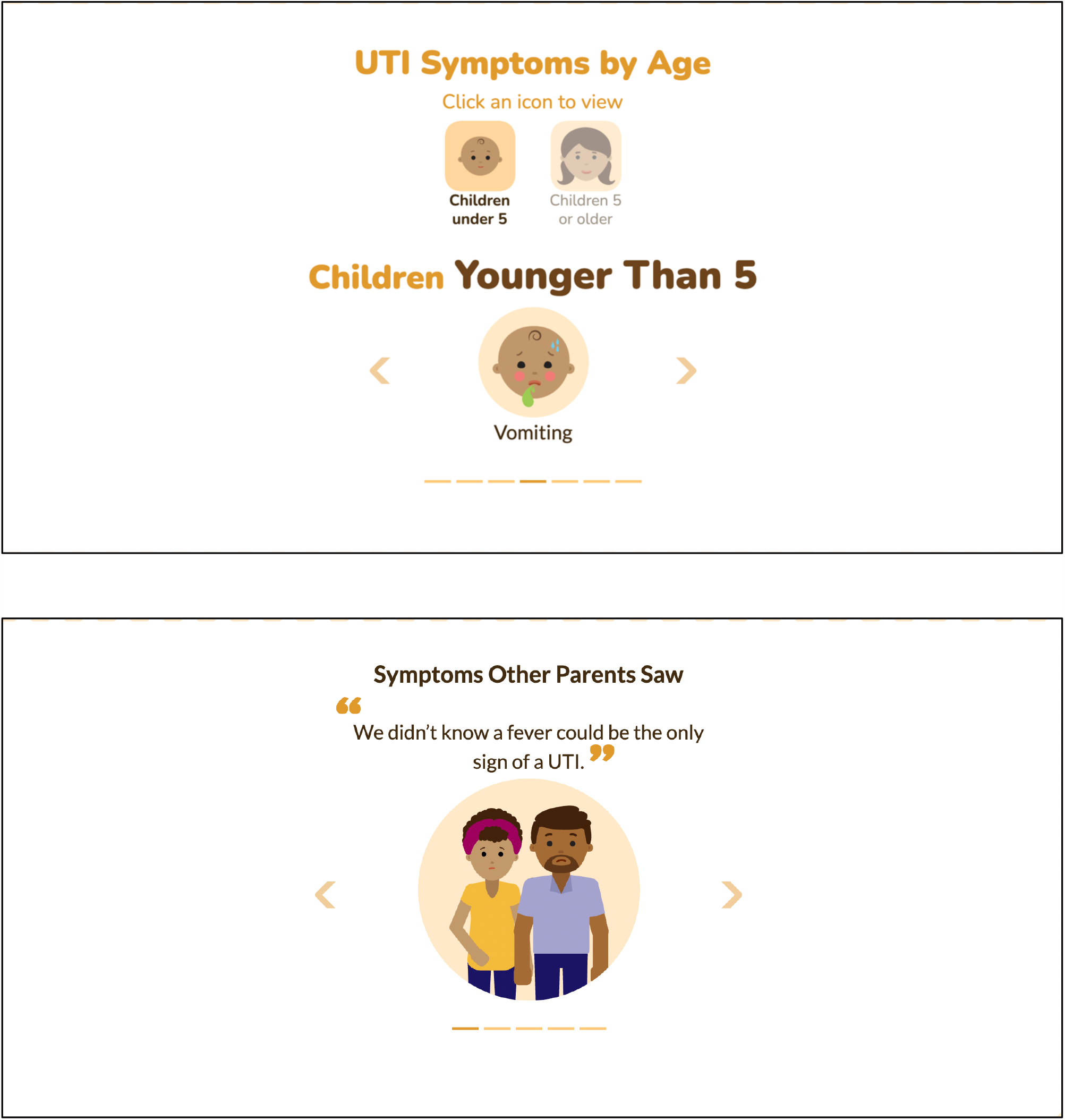

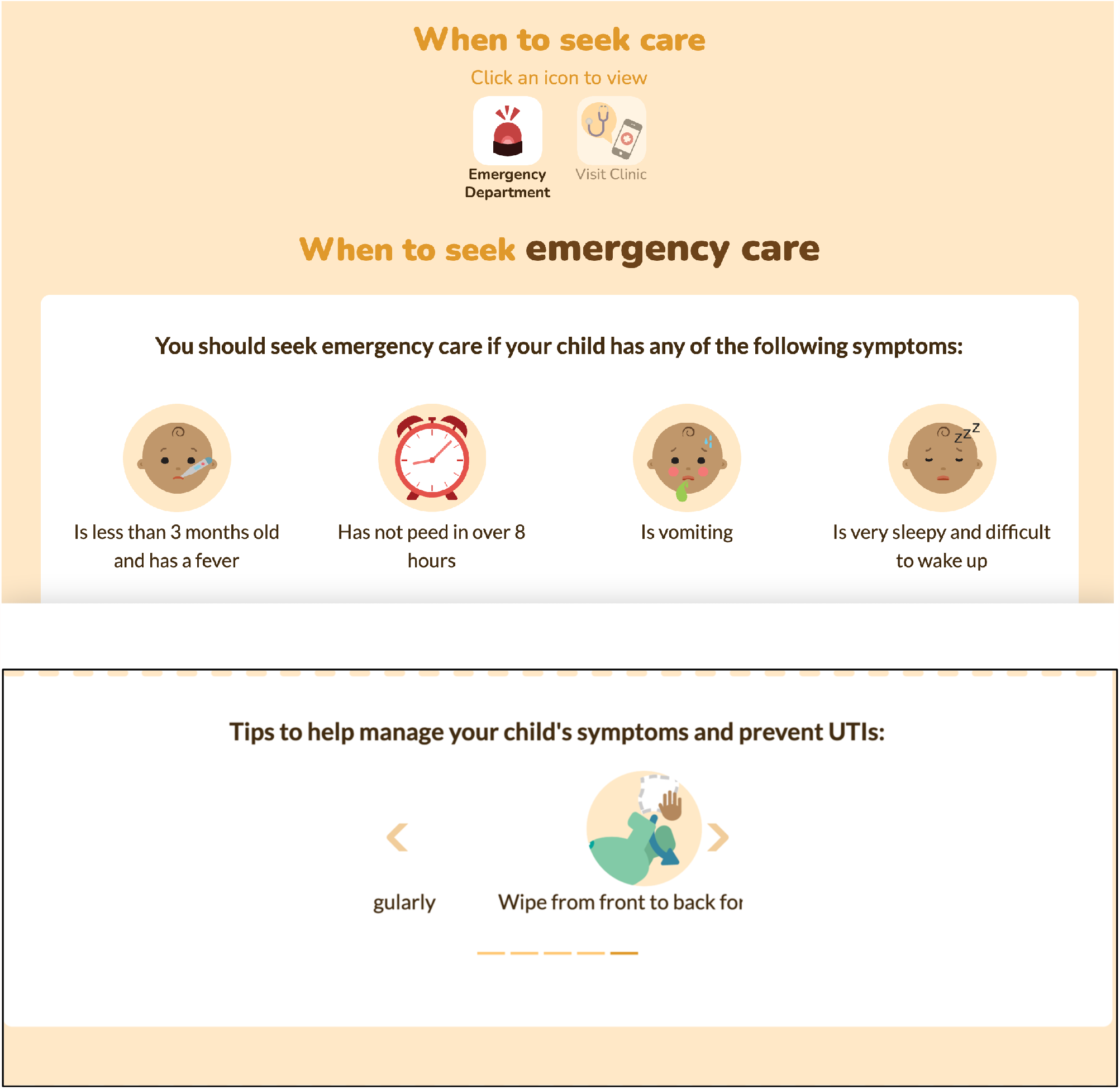

### Appendix D Usability Survey

#### SECTION 1: Demographics

1) What is your gender?
  □ Male
  □ Female
3) What is your Age?
  □ Less than 20 years old
  □ 20-30 years
  □ 31-40 years
  □ 41-50 years
  □ 51 years and older
4) What is your Marital Status?
  □ Married
  □ Single
5) What is your gross annual household income?
  □ Less than $25,000
  □ $25,000-$49,999
  □ $50,000-$74,999
  □ $75,000-$99,999
  □ $100,000-$149,999
  □ $150,000 and over
6) What is your highest level of education?
  □ Some high school
  □ High school diploma
  □ Some post-secondary
  □ Post-secondary certificate/diploma
  □ Post-secondary degree
  □ Graduate degree
  □ Other
7) How many children do you have?_______
8) How old are your children?_________

#### SECTION 2: Assessment of attributes of the arts-based, digital tools

***\*participant is randomized to view 1 of 2 digital tools then automatically directed to the survey***

1. It is useful. [5-point Likert Scale]
2. It provides information that is relevant to me as a parent. [5-point Likert Scale]
3. It is simple to use. [5-point Likert Scale]
4. I can use it without written instructions or additional help. [5-point Likert Scale]
5. Its length is appropriate. [5-point Likert Scale]
6. It is aesthetically pleasing (i.e., images, colours, etc.). [5-point Likert Scale]
7. It helps me to make decisions about my child’s health. [5-point Likert Scale]
8. I would use it in the future. [5-point Likert Scale]
9. I would recommend it to a friend. [5-point Likert Scale]
10. List the most negative aspects: [open text]
11. List the most positive aspects: [open text]

### Appendix E Project Timeline

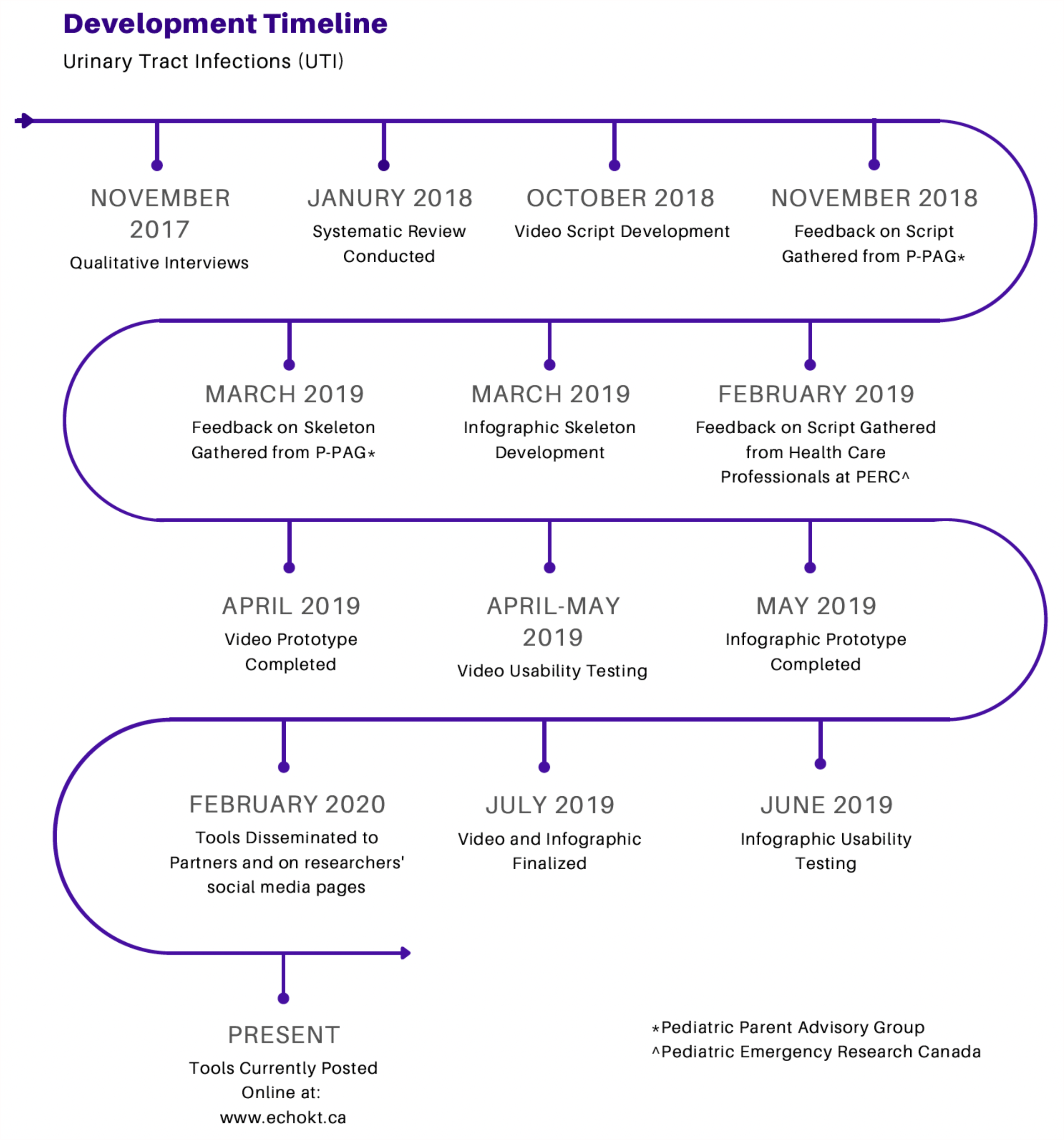

## Notes

### Competing Interest Statement

The authors have declared no competing interest.

### Funding Statement

This study was funded by the Networks of Centres of Excellence of Canada and the Women and Children's Health Research Institute

### Author Declarations

University of Alberta Research Ethics Boards (REB)

## Research Papers

Campbell, A., Hartling, L., Louie-Poon, S., Scott, S.D. (2021). Exploring the experiences and information needs of parents caring for a child with a urinary tract infection: A qualitative study. Journal of Patient Experience, 8, 1-8. https://doi.org/10.1177%2F23743735211008299.

Gates, A., Shulhan, J., Featherstone, R., Scott, S.D., Hartling, L. (2018) A systematic review of parents’ experiences and information needs related to their child’s urinary tract infection. Patient Education and Counseling, 101(7): 1207-1215. https://doi.org/10.1016/j.pec.2018.01.014.

## Presentations & Research Conferences

Sung, H., Le, A., Plesuk, T., Brooks, H.M., Ahn, T., Hartling, L., Scott, S.D. Usability of infographics for pediatric UTI and bronchiolitis. Shirley Stinson Nursing Research Conference. Edmonton, AB. November 7, 2019.

Plesuk, T., Louie-Poon, S., Campbell, A., Sung, H., Le, A., Brooks, H.M., Hartling, L., Scott, S.D. Comparing the usability evaluation results of two parental tools for UTI. Shirley Stinson Nursing Research Conference, Edmonton, AB. November 7, 2019.

Plesuk, T., Louie-Poon, S., Campbell, A., Sung, H., Le, A., Brooks, H.M., Hartling, L., Scott, S.D. Comparing the usability evaluation results of two parental tools for UTI. Women and Children’s Health Research Institute Research Day. Edmonton, AB. November 6, 2019.

Plesuk, T., Louie-Poon, S., Campbell, A., Sung, H., Le, A., Brooks, H.M., Hartling, L., Scott, S.D. Comparing the usability evaluation results of two parental tools for UTI. Women and Children’s Health Research Institute Public Lecture on Children’s Pain. Edmonton, AB. November 5, 2019.

Sung, H., Le, A., Plesuk, T., Brooks, H.M., Ahn, T., Hartling, L., Scott, S.D. Usability of infographics for pediatric UTI and bronchiolitis. Undergraduate Student Summer Series: Launchpad to Research, Edmonton, AB. July 24, 2019.

Gates, A., Shulhan, J., Featherstone, R., Scott, S.D., Hartling, L. (2017). A systematic review of parents’ self-reported experiences and information needs related to their child’s urinary tract infection. [oral presentation] Women & Children’s Health Research Institute Annual Research Day. Edmonton, Alberta. October 25, 2017.

## References

1. Kenney, K.M., Glynn, L.G., Dineen, B. (2010). A survey of the management of urinary tract infection in children in primary care and comparison with the NICE guidelines. BMC Family Practice, 11(6), 1–6. http://www.biomedcentral.com/1471-2296/11/6.

2. Montini, G., Tullus, K., Hewitt, I. (2011). Febrile urinary tract infections in children. New England Journal of Medicine, 365(3), 239–250.

3. Chang, S.L., Shortliffe, L.D. (2006). Pediatric urinary tract infections. Pediatr Clin N Am, 53, 379–400. https://doi.org/10.1016/j.pcl.2006.02.011.

4. Coulthard, M.G., Lambert, H.J., Vernon, S.J., Hunter, E.W., Keir, M.J., Matthews, J.N.S. (2014). Does prompt treatment of urinary tract infection in preschool children prevent renal scarring: Mixed retrospective and prospective audits. Arch Dis Child, 99, 342–347. https://doi.org/10.1136/archdischild-2013-304428

5. Shaikh, N., Morone, N.E., Bost, J.E., Farrell, M.H. (2008). Prevalence of urinary tract infection in childhood: A meta-analysis. Pediatr Infect Dis J, 27, 302–308. https://doi.org/10.1097/inf.0b013e31815e4122

6. Struthers, S., Scanlon, J., Parker, K., Goddard, J., Hallett, R. (2003). Parental reporting of smelly urine and urinary tract infection. Arch Dis Child, 88, 250–252. https://dx.doi.org/10.1136%2Fadc.88.3.250

7. Gates, A., Shulhan, J., Featherson, R., Scott, S.D., Hartling, L. (2018). A systematic review of parents’ experiences and information needs related to their child’s urinary tract infection. Patient Educ Couns, 101(7), 1207–1215. https://doi.org/10.1016/j.pec.2018.01.014

8. Harmsen, M., Wensing, M., van der Wouden, J.C., Grol, R.P.T.M. (2007). Parents’ awareness of and knowledge about young children’s urinary tract infections. Patient Education and Counseling, 66, 250–255. https://doi.org/10.1016/j.pec.2006.12.009

9. Owen, D., Vidal-Alaball, J., Mansour, M., Bordeaux, K., Verrier Jones, K., Edwards, A. (2003). Parents’ opinions on the diagnosis of children under 2 years of age with urinary tract infection. Family Practice, 20, 531–537. https://doi.org/10.1093/fampra/cmg507

10. Slater, M.D., Buller, D.B., Waters, E., Archibeque, M., LeBlanc, M. (2003). A test of conversational and testimonial messages versus didactic presentations of nutrition information. J Nutri Educ Behav, 35, 255–259. https://doi.org/10.1016/s1499-4046(06)60056-0

11. Scott, S.D., Osmond, M.H., O’Leary, K.A., Graham, I.D., Grimshaw, J., Klassen, T., and the Pediatric Emergency Research Canada MDI/spacer Study Group. (2009). Barriers and supports to implementation of MDI/spacer use in nine Canadian pediatric emergency departments: a qualitative study. Implementation Science, 4, 65. https://doi.org/10.1186/1748-5908-4-65

12. Scott, S.D., Brett-MacLean, P., Archibald, M., Hartling, L. (2013). Protocol for a systematic review of the use of narrative storytelling and visual-arts-based approaches as knowledge translation tools in healthcare. Systematic Reviews, 2, 19. https://doi.org/10.1186/2046-4053-2-19

13. Scott, S.D., Hartling, L., Klassen, T.P. (2009). The power of stories: Using narratives to communicate evidence to consumers. Evidence & Outcomes, 13(2), 109–111. https://doi.org/10.1111/j.1751-486x.2009.01401.x

14. Scott, S.D., Hartling, L., O’Leary, K., Archibald, M., Klassen, T. (2012). Stories – a novel approach to transfer complex health information to parents: A qualitative study. Arts & Health, 4(2), 162–173. https://doi.org/10.1080/17533015.2012.656203

15. Hartling, L., Scott, S.D., Pandya, R., Johnson, D., Bishop, T., Klassen, T.P. (2010). Storytelling as a communication tool for consumers: development of an intervention for parents of children with croup. Stories to communicate health information. BMC Pediatrics, 10, 64. http://www.biomedcentral.com/1471-2431/10/64

16. Hartling, L., Scott, S.D., Johnson, D.W., Bishop, T., Klassen, T.P. (2013). A randomized controlled trial of storytelling as a communication tool. PLOS, 8(10), e77800. https://doi.org/10.1371/journal.pone.0077800

17. Gates, A., Shulhan, J., Featherstone, R., Scott, S.D., Hartling, L. (2018) A systematic review of parents’ experiences and information needs related to their child’s urinary tract infection. Patient Education and Counseling, 101(7): 1207–1215. https://doi.org/10.1016/j.pec.2018.01.014.

18. Campbell, A., Hartling, L., Louie-Poon, S., Scott, S.D. (2021). Exploring the experiences and information needs of parents caring for a child with a urinary tract infection: A qualitative study. Journal of Patient Experience, 8, 1–8. https://doi.org/10.1177%2F23743735211008299.

19. TRanslating Emergency Knowledge for Kids (TREKK). UTI – Bottom Line Recommendations. July 2019 – Version 1.0. Retrieved from: https://trekk.ca/resources?external_resource_type=Quick_glance&tag_id=D014552

20. Hornbæk, K. (2006). Current practice in measuring usability: Challenges to usability studies andresearch. International journal of human-computer studies. 64(2), 79–102.

21. Carifio, J., & Perla, R. (2008). Resolving the 50-year debate around using and misusing Likert scales. Blackwell Publishing, 42, 1150–1152.

22. Sullivan, G.M., & Artino Jr., A.R. (2013). Analyzing and interpreting data from Likert-type scales. Journal of Graduate Medical Education, Editorial.

23. Featherstone, R.M., Leggett, C., Knisley, L., Jabbour, M., Klassen, T.P., Scott, S.D., Van De Mosselaer, G., Hartling, L. (2018). Creation of an integrated knowledge translation process to improve pediatric emergency care in Canada. Health Commun, 33(8), 980–987. https://www.doi.org/10.1080/10410236.2017.1323538.

